# Efficacy of premarital genotype screening and counselling on knowledge toward Sickle Cell disease among university students in Dodoma Tanzania: uncotrolled quasi-experimental study

**DOI:** 10.1101/2022.04.11.22273743

**Authors:** Arnold Gideon Lumbe, Stephen M Kibusi

## Abstract

**Background:** Tanzania is experiencing the increase burden of Sickle cell disease, with an estimate of 20.6% prevalence Sickle Cell carriers. However, there is no preventive measure has been put in this area by the government; a great focus has been directed in the diagnosis and management and national guideline emphasis on the care of people affected by Sickle Cell Disease.

**Methods:** A non-controlled quasi-experimental study was conducted from June to September 2020 among 697 randomly recruited first-years university students from the University of Dodoma. Pre and post-test knowledge information were collected through structured self-administered questionnaires. Data was analyzed using SPSS v20. Simple and multiple linear analysis model was used to test for significant association of variables at 95% CI, at p<0.05. The results were presented using tables and figures.

**Results:** The mean knowledge score at pretest was 0.009±1.014 which improved to 0.365±0.901 on the posttest, with a statistically significant difference (t=6.965, p<0.01). The results of linear regression showed that knowledge change was not statistically associated with other predictors (p>0.05)

**Conclusion:** Health education demonstrated to be effective towards change in knowledge on sickle cell disease among University students.

## Background

Sickle Cell Disease (SCD) is one of the biggest health challenges worldwide (WHO, 2017). It is a group of inherited red blood disorders transmitted from parents to their offspring. Normally, healthy red blood cells are round and they move through small blood capillaries carrying oxygen to all parts of the body (Ghimire, 2016). But in individual affected with SCD, the red blood cells become hard and adhesive and appear like a C-shaped farm tool termed a “sickle”. As a result these abnormal Sickle cells die early within 10–17 days in contrast to the normal 120-day lifespan of non-sickled RBCs, leading to constant shortage of red blood cells (Babalola et al., 2019). Sickle cells can get jammed in small blood vessels and block the flow of blood and oxygen to organs in the body. These blockages cause repeated episodes of severe pain, organ damage, serious infections, or even stroke (Aderotoye-oni, Diaku-akinwumi, Adeniran, & Falase, 2018).

Regardless of the increase in the disease, diagnosis, and understanding causes of the inherited disorders, thousands of children die through the lack of preventive measures, like premarital Sickle Cell genotype screening by two intending married individuals to identify their genotype before marriage (WHO, 2018).

According to WHO (2018), 300,000 children with SCD are born worldwide every year. Moreover, about 5% of the population in the world carries hemoglobinopathy genes that cause SCD (WHO, 2018). Most of the affected people are those coming from sub-Saharan Africa, Saudi, India, Arabia, and the Mediterranean of which carries 75% of all SCD disease (WHO, 2017).

There are 275,000 babies in Africa born with SCD annually. In the absence of adequate preventive intervention, this projection is expected to reach 400,000 by 2050 (WHO, 2017). This increase in affected people is compounded by 6.4% of under-five mortality in Africa due to SCD (Faremi, 2018). The most affected countries are Nigeria with a prevalence of SCT 24%, SCD between 2 to 3% (Kambasu, Rujumba, Lekuya, Munube, & Mupere, 2019), Ghana, 24–30 % of the population carries the SCT and 2% has SCD (Asare et al., 2018). The Democratic Republic of Congo, SCT prevalence is estimated to be 18.3% and SCD at 1.4% (WHO, 2019).

Despite that many people are affected by SCD, the level of knowledge on SCD remains low. Various studies have revealed low knowledge of SCD among university students (Asare et al., 2018; Boadu, 2018; Mombo et al., 2017; Salman & Abass, 2019). Thus, there is a need to improve SCD knowledge to break the Sickle Cell disease cycle in the population. Improved knowledge towards premarital screening and counseling are some of the principal approaches to achieve this purpose (Al-Balushi & Al-Hinai, 2018).

Another possible intervention to make is genetic counseling. In this, a person is informed about their genetic predisposition to diseases. Couples need to be aware of the possible genetic features of their unborn children (Hussaini, Durbunde, Jobbi, Muhammad, & Mansur, 2019). This, along with premarital screening can help in achieving the desired level of knowledge, in promoting health and improving the quality of life of young people who are about to start planning for long-term relationships and marriage (Ango et al., 2018). Knowledge about screening premarital Sickle Cell genotype can be obtained through training programmes (Lola et al., 2018; Review & Bridget, 2015).

Since university life is complex and interactive, some students can start the marital engagement or even become mothers and fathers while at college (Chiao, Yi, & Ksobiech, 2012; Omuemu, Obariagbon, & Ogboghodo, 2012). This is a reason why some studies support the assessment of university students’ knowledge of premarital screening and counseling as they continue with their studies(Abioye-Kuteyi, Oyegbade, Bello, & Osakwe, 2018; Faremi & Olatubi, 2018b).

Tanzania is one of the countries that have high annual births of Sickle Cell disease worldwide (WHO Afica, 2018). The prevalence of Sickle Cell carriers (HbAS) is 20.6% (MoHCDGEC, 2020). The statistics show that there 6 in every 1000 children born with SCD annually (Makani et al., 2018). This contributes to up to 7% of total deaths of the under-five children, if this situation continues without intervention, up to 90% of the affected children are predicted not to survive beyond childhood (MoHCDGEC, 2016).

What gives hope is that the problem can be contained if timely and appropriate intervention is made. There is evidence that increased awareness and knowledge to the community, especially the youth on premarital Sickle Cell genotype screening and counseling work better toward preventing SCD and, hence, alleviate the impact on the general population (WHO, 2017). While this observation if fortiori, there is limited information on the level of knowledge among the youth in Tanzania. This information is required for appropriate interventions, especially among the university students who most of them are observed to get married in the course of their studies at university. However, studies from other areas of Africa reported that university students are a good target to imparting knowledge on premarital Sickle Cell genotype screening and counseling (Hussaini et al., 2019; Salman & Abass, 2019).

Little attention has been put in this area by the government and stakeholders; however, a great focus has been directed in the diagnosis and management of SCD and national guideline emphasis on the care of people affected by SCD (Ambrose et al., 2018; Makani et al., 2015). Therefore, this study attempted to fill this gap by testing the efficacy of premarital genotype screening and counseling on knowledge toward SCD among university students in Tanzania.

## Methods

### Study Area and Design

Uncontrolled quasi-experimental (pre and post-test only) coupled with the quantitative approach was conducted in Dodoma Region from June to September 2020 involving the university students from the University of Dodoma. Current the university enrollment rate is around 30,000 students, which is 80% of its capacity to enroll (UDOM 2019). Moreover, the university has seven colleges, namely the College of Earth Science, College of Natural and Mathematical Sciences, College of Informatics and Virtual Education, College of Education, College of Humanities and Social Sciences, College of Health and Allied Science. It is a big university in central and east Africa, which occupies students from all regions of Tanzania.

#### Study Population

The research included all unmarried first-year bachelor degree students at the University of Dodoma, sampled from different degree programmes offered at the University.

#### Inclusion and Exclusion Criteria

The research included first-year bachelor degree students who were perusing different degree programmes, aged between 19 to 29 years, consented to participate in the study, all unmarried and not known to have Sickle Cell disease. The study excluded all pregnant female students, students who were severely ill. Likewise, all first-year students who had repeated the course were excluded in the study.

#### Sample Size Determination and Sampling Technique

The sample size for the pre and posttest group was computed by using A Cochran formula (1977), whereby 95% confidence level,19.7 mean knowledge score at posttest with 3.1 statistical significant difference with normal standard deviation of 0.84. The estimated total sample size was 697 participants.

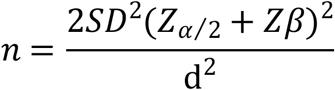

##### Sampling techniques

Multistage sampling technique was used to select the required subjects for the study. The procedure followed four stages as shown below:

##### Stage one

Six colleges which were not providing health and allied sciences programmes from the University of Dodoma were selected purposively.

##### Stage two

By average, every collage comprises about 6 bachelor degree programmes giving a total of 45 programmes. Out of this, 18 programmes were selected. A simple random technique employed to select 3 bachelor programs per college using a replacement lottery method.

##### Stage three

Each selected programme was categorized as a stratum. The stratified sampling techniques were employed to get the proportional allocation of participants from every programme selected.

##### Stage four

The study participants were selected using a systematic sampling technique after calculating the k^th^ to get the interval of picking the eligible participant for the study.

#### Data Collection Procedure

The data were collected using a structured questionnaire adopted and modified to fit for the current study. The questionnaire before it was employed for data collection, its internal consistency measured 0.908. The questionnaire was also pretested involving 64 students from St.Johns’ University of Tanzania.

#### Data Collection Techniques

Data were collected from 5^th^ June to 10^th^ September 2020. Before data collection; the researcher employed purposively 6 researcher assistants who were 4 nurses, their responsibility was to deliver health education and counseling, and 2 laboratory technicians who employed for screening purpose. They were trained to deliver health information, use questionnaire. The data was collected in three stages; named, pre-test, health education intervention, and post-test. The details of each phase are given hereunder:

#### Pre-test Intervention Stage

This involved the collection of baseline data from the students through a self-administered questionnaire. In this stage, the researcher categorized participants into six groups based on their respective colleges. Then, six lecture rooms were selected from each college which was used by the study participants during the first stage of data collection “pretest”. The procedure started by rapport building by introducing each other to participants and briefly explained the nature and the purpose of the study. After that, the researcher and research assistant provided an overview of the purpose of the study and the assessment tool which will be used during data collection. Then, the participants were given a consent form to seek either to accept or refuse to participate in the study. After obtaining the consent, self-administered questionnaires were distributed to each study participants, to obtain their social-demographic information, baseline knowledge, and attitude toward Sickle Cell disease. The whole process took 30 minutes, and then questionnaires were collected for initial analysis, to identify baseline knowledge.

#### The Health Education Stage

This was the interventional stage at which health education were conducted, the whole intervention process took 8weeks. This procedure was done into the participants’ respective colleges. The participants were divided into 11 groups comprised of 53 participants each. Lectures and discussions method were used to deliver materials to participants. The audio-visual equipment like the computer and LCD projector were used to facilitate effective communication. The sessions used Sickle Cell teaching package adopted from WHO (2006). The contents of the package comprised the definition of Sickle Cell disease, causes of the disease, signs, and symptoms, complications of SCD, prevention through premarital screening. The time allocated for sessions was 45 minutes for lecture discussion. The participants were given take-home reading hand handouts to keep them revising contents taught in the class.

#### Post Intervention Stage

This was performed on the 8^th^ week after implemented health education and screening for the same participants. The researcher communicated with group representatives through phone to inform the participants to be in the same venue used during pre-test. The same questionnaires were used to obtain a post-intervention level of knowledge on Sickle Cell disease. The researcher and research assistant were divided into six previous group used in pre-test to participants’ respective colleges, whereby they re-introduced and explain the purpose of the second round data collection. Then, questionnaires were distributed to participants, after 25 minutes it was completed filled and were collected for data analysis. This stage marked the end of data collection; the obtained data were subjected to analysis.

### Validity and Reliability

#### Validity

According to Parahoo (2006, p. 300–309), validity is classified in three forms namely; content, construct, and criterion. Content validity of questionnaires means an item of questionnaire represents the phenomenon being studied; Construct validity of questionnaires is the ability of the questionnaire to measure what is intended; and criterion validity is the extent to which a measure is related to an outcome. To achieve the validity of the instrument, the researcher used the standardized tool adopted from previous research with Chronbach’s Alpha of 0.908 indicating content acceptance for its usefulness. Moreover, content validity was observed by formulating an adequate sample of the question for the variables included in the topic under study, basing on the objectives that measured the intended outcome. Moreover, the construct validity of the questionnaire was assured by careful formulating questionnaire after a thorough theoretical and research literature review on the topic under study was done to get in-depth knowledge of formulating data collection tools.

#### Reliability

To ensure reliability in this research, the researcher did the following: first six research assistant were trained on the use of the questionnaire, use of SCD content to train participants and counselling to participants found to have traits of Sickle Cell. Training took one week and research assistants were given a chance to practice training before they went to intervene. To ensure consistency of the variables included in the study, data collection tools were pre-tested to 64 students at St. Johns University of Tanzania. The participants who were used in the pre-test were not involved in the actual study. Minor correction for typographic errors and clarity was done.

### Measurement of Variables

#### Covariate variable

Demographic information was considered a covariate variable. They were measured by 17 questions. Age of the respondent was measured in a continuous variable while place of residence, program of study, tribe, sex, religion, college, parents education, type of family and history of Sickle Cell disease was measured in the categorical variable.

#### Knowledge

Knowledge on sickle cell disease was measured by using 7 domains with 5 sub-domain each including;concept of SCD, causes of SCD, Screening, management, complication, preventive measures and benefits of SCD screening.The response of each item was yes/no and not sure answers, which were then converted into correct and incorrect. The principal component analysis (PCA) was done to determine the mean scores that determined the cut-point for individual knowledge level (Kotb *et al*., 2019). Percentile was used as a cutoff point to categorize the level of knowledge in low level of knowledge, moderate and high level knowledge.

#### Data Analysis

The Statistical Package for Social Science (SPSS) software version 20 developed by IBM Corporation in 2015 was used for data entry, coding, scoring, and analysis. Descriptive statistics analysis was done to present the frequency and percentages of data consisting of social demographic and prevalence of Sickle Cell Disease (SCD status). Graphical presentation was used for illustrations such as frequency distribution table, histogram. Moreover, Chi-square was used for categorical data to determine the factors related to the outcome variables. In addition to that, paired sample t-test analysis was employed to determine the difference between pre and post-intervention sample mean scores for outcome variables.

## Results

### Socio-demographic characteristics of the study participants

Six hundred and ninety seven participants involveld in the study. Majority of the respondents 549 (78.8%) were aged 19 to 29 years, with a mean (SD) age of 23.20 ± 2.00 years. By gender composition, more than half of participants 416 (59.7%) were male.With regard of residence most of participants 469(67.3 %) were from rural and about 216 (31.0 %) had come from lake zones. On respect to religion, 579 (83.1%) were Christian. Regarding history of sickle cell to participants family; only 110 (15.8%) reported to have sickle cell to their family.About head of the family of the participants, majorital of them 592 (84.9%) headed by their father.On respect to media of communication majority 691 (99.1%) used phones as a sourse of getting information. Findings of other socio-demographic characteristic are found in (Table 1).

**Table 1:** Social-demographic Characteristics of the Study Participants (n = 697)

| **Variable** | **Frequency**<br>**(N)** | **Percentage**<br>**(%)** |
| --- | --- | --- |
| Age |  |  |
| Mean (M) 23.20 |  |  |
| Minimum 19 years |  |  |
| Maximum 29 years |  |  |
| 19 to 24 years | 549 | 78.8 |
| 25 to 29 years | 148 | 21.2 |
| Sex |  |  |
| Male | 416 | 59.7 |
| Female | 281 | 40.3 |
| Zone distribution of participants |  |  |
| Central zone | 92 | 13.2 |
| Coastal zone | 102 | 14.6 |
| Lake zone | 216 | 31 |
| Northern zone | 122 | 17.5 |
| South highland zone | 126 | 18.1 |
| Western zone | 25 | 3.6 |
| Zanzibar | 14 | 2 |
| Residence group of participants |  |  |
| Rural | 469 | 67.3 |
| Urban | 228 | 32.7 |
| Religion of participants |  |  |
| Christians | 579 | 83.1 |
| Muslim | 118 | 16.9 |
| Occupation status of participants |  |  |
| Employed |  |  |
| Yes | 19 | 2.7 |
| No | 678 | 97.3 |
| College of participants |  |  |
| COED | 159 | 22.8 |
| CIVE | 103 | 14.8 |
| CNMS | 52 | 7.5 |
| CBSL | 145 | 20.8 |
| COES | 127 | 18.2 |
| CHSS | 111 | 15.9 |
| Type of media used by participants to get information |  |  |
| Phone |  |  |
| Yes | 691 | 99.1 |
| No | 6 | 0.9 |
| Magazine |  |  |
| Yes | 49 | 7 |
| No | 648 | 93 |
| Television |  |  |
| Yes | 113 | 16.2 |
| No | 584 | 83.8 |
| Total number of media used |  |  |
| 1 media | 561 | 80.5 |
| 2media | 101 | 14.5 |
| >2 media | 35 | 5 |
| Type of participants' family |  |  |
| Nuclear | 539 | 77.3 |
| Extended | 158 | 22.7 |
| Head of the family |  |  |
| Father | 592 | 84.9 |
| Mother | 90 | 12.9 |
| Relatives | 15 | 2.2 |
| History of Sickle Cell disease in participants' family |  |  |
| Yes | 110 | 15.8 |
| No | 587 | 84.2 |

### Level of knowledge responses in pre and posttest items analysis of sickle cell disease among university students in Dodoma region

About 16 questions asked on knowledge of Sickle Cell disease. The responses were yes, no and not sure, and then converted into correct/incorrect responses. The findings on item asked Sickle Cell disease-cause red blood cell to become sickle-shaped and hard, the proportion of participants responded correctly were 143 (20.5%) at pre-test, after health education most of them at post-test 423 (60.7%) responded correct. About causes of SCD on item asked, when both parents have SCD trait, chance of each pregnant having a child with Sickle Cell disease is-25%,the proportion of particiant responded correctly at pre-test were 200 (28.7%) while after health education most of them 572(82.1%) at post-test answered correctly. Moreover, about prevention domain at pre-test few of them 182 (26.1%) answered correctly on item asked the best way to prevent Sickle Cell disease is two persons with SCD not marrying each other, while at post-test most of them 471(67.6%) responded correctly. More details about the Level of knowledge responses in pre and posttest items analysis of sickle cell disease among university students in Dodoma region are as shown in the table 2.

**Table 2:**
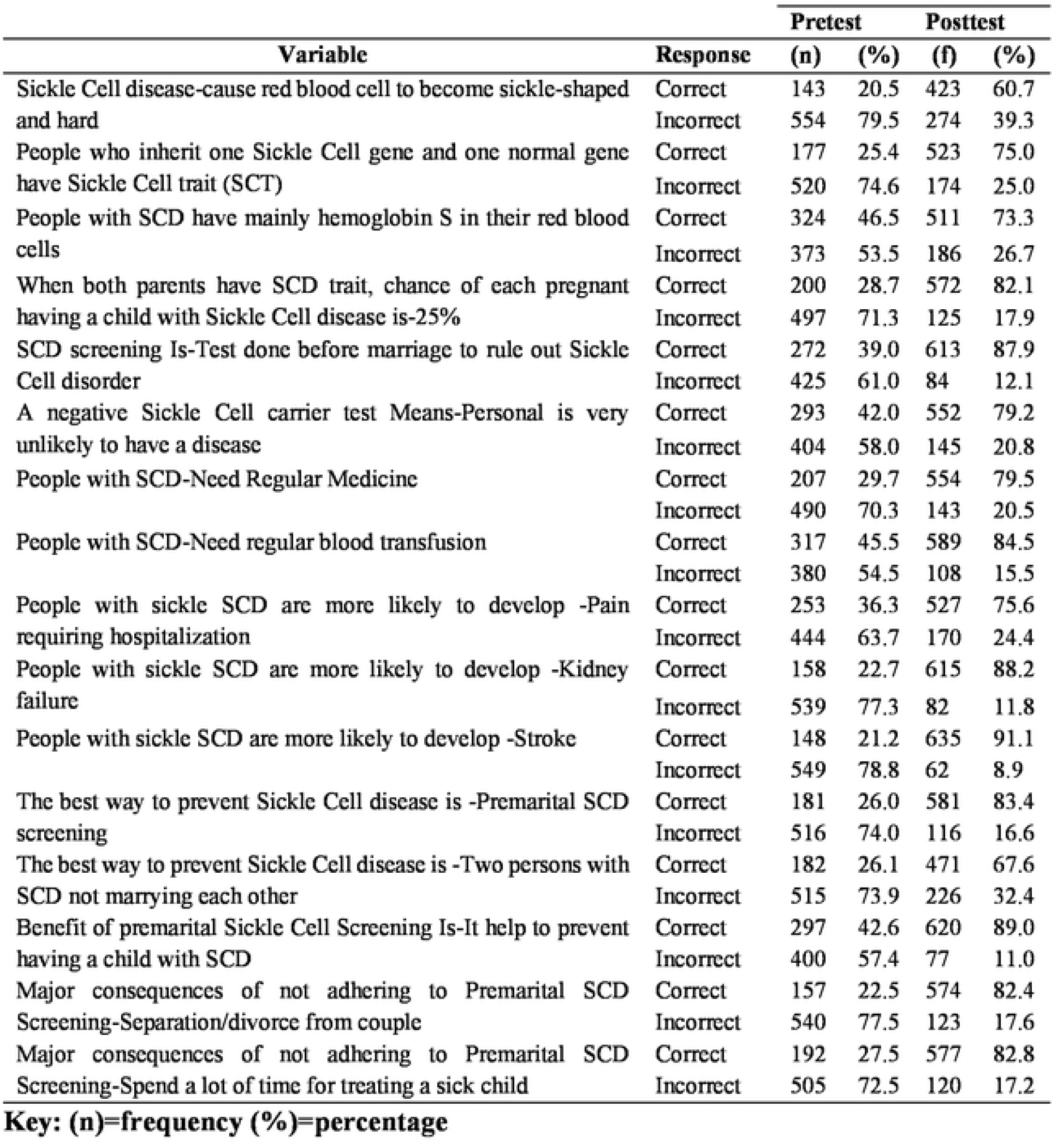
Level of knowledge responses in pre and post-test items analysis of sickle cell disease among university students in Dodoma region (n=697)

| Variable | Response | Pretest |  | Posttest |  |
| --- | --- | --- | --- | --- | --- |
|  |  | (n) | (%) | (f) | (%) |
| Sickle Cell disease-cause red blood cell to become sickle-shaped and hard | Correct | 143 | 20.5 | 423 | 60.7 |
|  | Incorrect | 554 | 79.5 | 274 | 39.3 |
| People who inherit one Sickle Cell gene and one normal gene have Sickle Cell trait (SCT) | Correct | 177 | 25.4 | 523 | 75.0 |
|  | Incorrect | 520 | 74.6 | 174 | 25.0 |
| People with SCD have mainly hemoglobin S in their red blood cells | Correct | 324 | 46.5 | 511 | 73.3 |
|  | Incorrect | 373 | 53.5 | 186 | 26.7 |
| When both parents have SCD trait, chance of each pregnant having a child with Sickle Cell disease is-25% | Correct | 200 | 28.7 | 572 | 82.1 |
|  | Incorrect | 497 | 71.3 | 125 | 17.9 |
| SCD screening Is-Test done before marriage to rule out Sickle Cell disorder | Correct | 272 | 39.0 | 613 | 87.9 |
|  | Incorrect | 425 | 61.0 | 84 | 12.1 |
| A negative Sickle Cell carrier test Means-Personal is very unlikely to have a disease | Correct | 293 | 42.0 | 552 | 79.2 |
|  | Incorrect | 404 | 58.0 | 145 | 20.8 |
| People with SCD-Need Regular Medicine | Correct | 207 | 29.7 | 554 | 79.5 |
|  | Incorrect | 490 | 70.3 | 143 | 20.5 |
| People with SCD-Need regular blood transfusion | Correct | 317 | 45.5 | 589 | 84.5 |
|  | Incorrect | 380 | 54.5 | 108 | 15.5 |
| People with sickle SCD are more likely to develop -Pain requiring hospitalization | Correct | 253 | 36.3 | 527 | 75.6 |
|  | Incorrect | 444 | 63.7 | 170 | 24.4 |
| People with sickle SCD are more likely to develop -Kidney failure | Correct | 158 | 22.7 | 615 | 88.2 |
|  | Incorrect | 539 | 77.3 | 82 | 11.8 |
| People with sickle SCD are more likely to develop -Stroke | Correct | 148 | 21.2 | 635 | 91.1 |
|  | Incorrect | 549 | 78.8 | 62 | 8.9 |
| The best way to prevent Sickle Cell disease is -Premarital SCD screening | Correct | 181 | 26.0 | 581 | 83.4 |
|  | Incorrect | 516 | 74.0 | 116 | 16.6 |
| The best way to prevent Sickle Cell disease is -Two persons with SCD not marrying each other | Correct | 182 | 26.1 | 471 | 67.6 |
|  | Incorrect | 515 | 73.9 | 226 | 32.4 |
| Benefit of premarital Sickle Cell Screening Is-It help to prevent having a child with SCD | Correct | 297 | 42.6 | 620 | 89.0 |
|  | Incorrect | 400 | 57.4 | 77 | 11.0 |
| Major consequences of not adhering to Premarital SCD Screening-Separation/divorce from couple | Correct | 157 | 22.5 | 574 | 82.4 |
|  | Incorrect | 540 | 77.5 | 123 | 17.6 |
| Major consequences of not adhering to Premarital SCD Screening-Spend a lot of time for treating a sick child | Correct | 192 | 27.5 | 577 | 82.8 |
|  | Incorrect | 505 | 72.5 | 120 | 17.2 |

### Overall level of knowledge on sickle cell disease among university students in Dodoma Region

Knowledge score index of participants was categorized by use of percentiles, described into 25, 50, 75 and 100 percentiles. Score index was categorized as 0 – 49 had low level of knowledge, 50 – 74 had moderate level of knowledge and 75 – 100 had high level of knowledge. The results indicated that at pre-test, about half of participants 356 (51.1%) had low level of knowledge, while 168 (24.1%) had moderate level of knowledge and few 173 (24.8%) had high level of knowledge. The knowledge score increased at post test posttest of which majority 467(67%) had high level of knowledge, some of them 119 (17%) had moderate level of knowledge and few111 (16 %) had low level of knowledge (Figure 7).

### Mean score differences of knowledge on Sickle Cell disease in pre and post-test among university students in Dodoma Region

A comparative analysis by paired t-test was performed to determine pre and posttest mean score differences of knowledge on Sickle Cell disease among university students. Findings in table 3, reveled there was a statistical significant means score differences of overall knowledge about premarital Sickle Cell disease between pretest (M=0.009; SD 1.014) and posttest (M=0.365; SD 0.930) with mean score difference (MD=0.365; SD=1.384) [t (696) =6.965; p <0.01; 95% CI: 0.262, 0.468)].

**Table 3:**
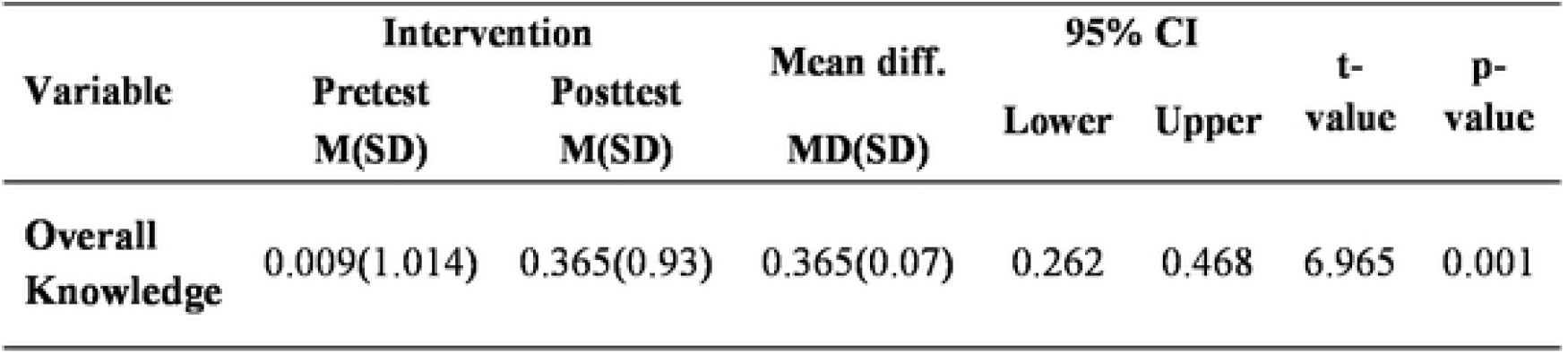
Mean score differences of knowledge on Sickle Cell disease in pre and post-test among university students in Dodoma Region (n=697)

### Simple linear regression on predictors of level of knowledge change towards Sickle Cell disease among university students in Dodoma region

The level of knowledge changes as dependent variable was analyzed associating for other predictors.The findings indicates predictors did not significantly associated with knowledge change (p>0.05) (Table 4).

**Table 4:**
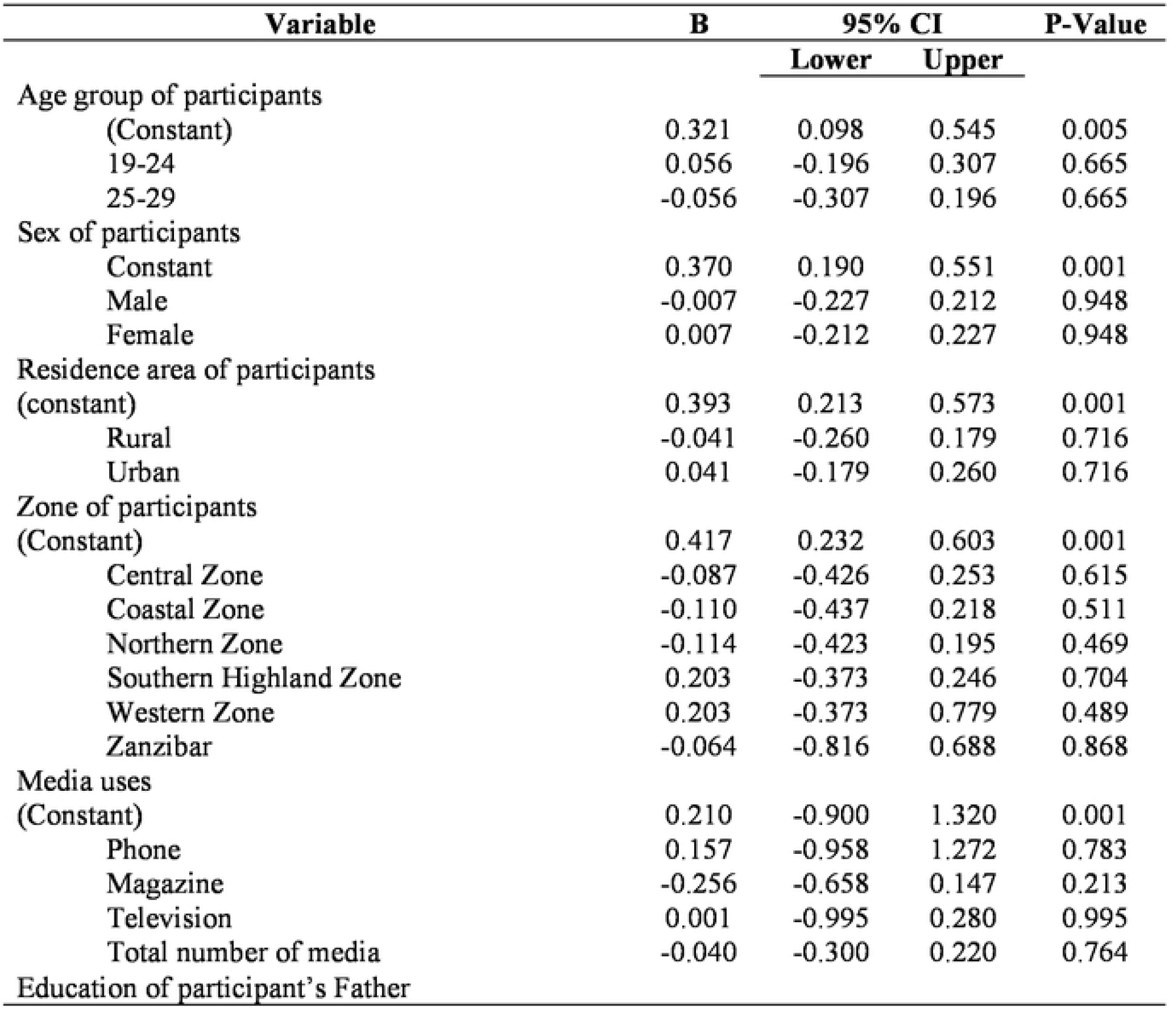
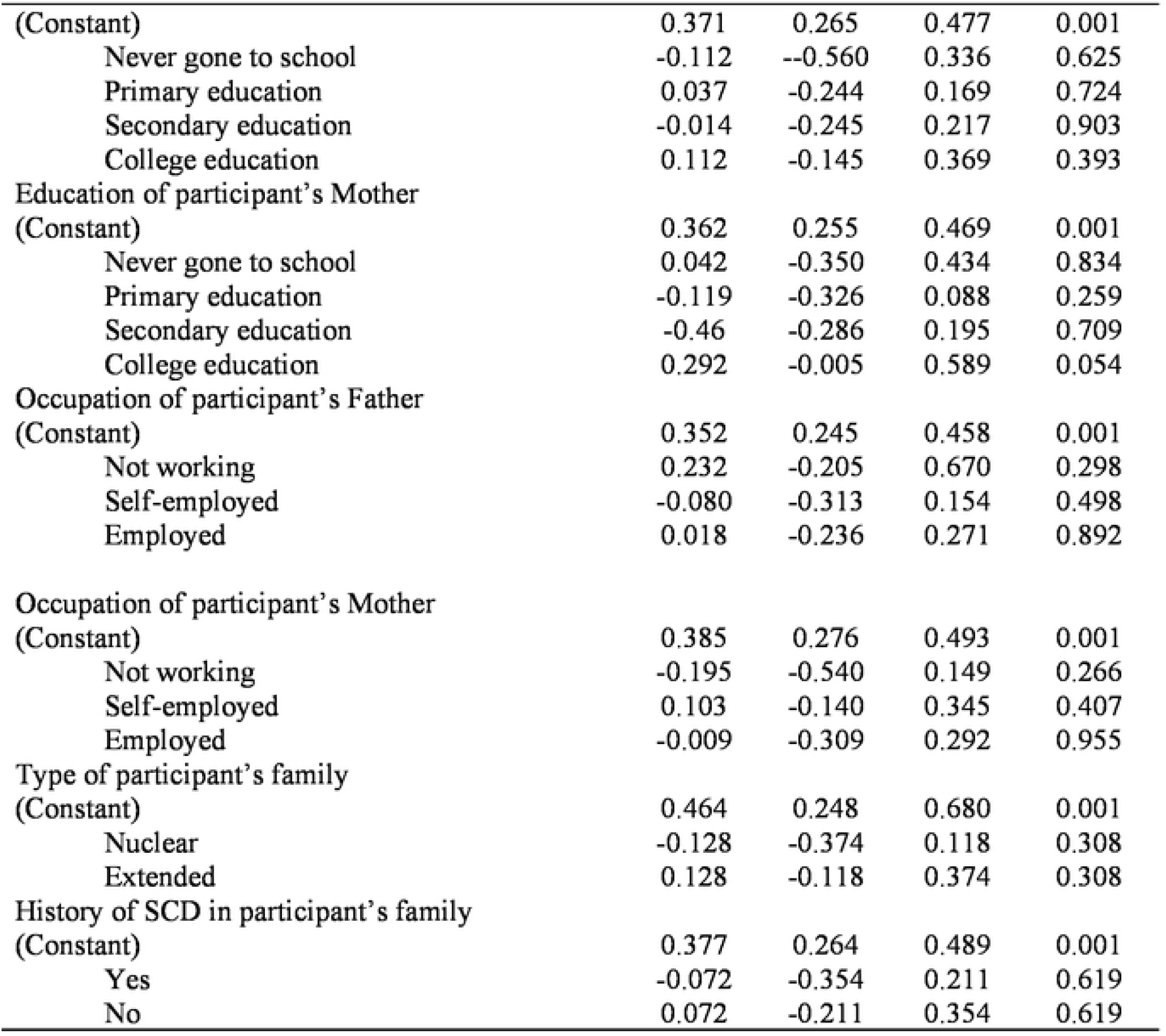
Simple linear regression on predictors of level of knowledge change towards Sickle Cell disease among university students(n-697)

| Variable | B | 95% CI |  | P-Value |
| --- | --- | --- | --- | --- |
|  |  | Lower | Upper |  |
| Age group of participants |  |  |  |  |
| (Constant) | 0.321 | 0.098 | 0.545 | 0.005 |
| 19-24 | 0.056 | -0.196 | 0.307 | 0.665 |
| 25-29 | -0.056 | -0.307 | 0.196 | 0.665 |
| Sex of participants |  |  |  |  |
| Constant | 0.370 | 0.190 | 0.551 | 0.001 |
| Male | -0.007 | -0.227 | 0.212 | 0.948 |
| Female | 0.007 | -0.212 | 0.227 | 0.948 |
| Residence area of participants |  |  |  |  |
| (constant) | 0.393 | 0.213 | 0.573 | 0.001 |
| Rural | -0.041 | -0.260 | 0.179 | 0.716 |
| Urban | 0.041 | -0.179 | 0.260 | 0.716 |
| Zone of participants |  |  |  |  |
| (Constant) | 0.417 | 0.232 | 0.603 | 0.001 |
| Central Zone | -0.087 | -0.426 | 0.253 | 0.615 |
| Coastal Zone | -0.110 | -0.437 | 0.218 | 0.511 |
| Northern Zone | -0.114 | -0.423 | 0.195 | 0.469 |
| Southern Highland Zone | 0.203 | -0.373 | 0.246 | 0.704 |
| Western Zone | 0.203 | -0.373 | 0.779 | 0.489 |
| Zanzibar | -0.064 | -0.816 | 0.688 | 0.868 |
| Media uses |  |  |  |  |
| (Constant) | 0.210 | -0.900 | 1.320 | 0.001 |
| Phone | 0.157 | -0.958 | 1.272 | 0.783 |
| Magazine | -0.256 | -0.658 | 0.147 | 0.213 |
| Television | 0.001 | -0.995 | 0.280 | 0.995 |
| Total number of media | -0.040 | -0.300 | 0.220 | 0.764 |
| Education of participant's Father |  |  |  |  |
| (Constant) | 0.371 | 0.265 | 0.477 | 0.001 |
| Never gone to school | -0.112 | -0.560 | 0.336 | 0.625 |
| Primary education | 0.037 | -0.244 | 0.169 | 0.724 |
| Secondary education | -0.014 | -0.245 | 0.217 | 0.903 |
| College education | 0.112 | -0.145 | 0.369 | 0.393 |
| Education of participant's Mother |  |  |  |  |
| (Constant) | 0.362 | 0.255 | 0.469 | 0.001 |
| Never gone to school | 0.042 | -0.350 | 0.434 | 0.834 |
| Primary education | -0.119 | -0.326 | 0.088 | 0.259 |
| Secondary education | -0.46 | -0.286 | 0.195 | 0.709 |
| College education | 0.292 | -0.005 | 0.589 | 0.054 |
| Occupation of participant's Father |  |  |  |  |
| (Constant) | 0.352 | 0.245 | 0.458 | 0.001 |
| Not working | 0.232 | -0.205 | 0.670 | 0.298 |
| Self-employed | -0.080 | -0.313 | 0.154 | 0.498 |
| Employed | 0.018 | -0.236 | 0.271 | 0.892 |
| Occupation of participant's Mother |  |  |  |  |
| (Constant) | 0.385 | 0.276 | 0.493 | 0.001 |
| Not working | -0.195 | -0.540 | 0.149 | 0.266 |
| Self-employed | 0.103 | -0.140 | 0.345 | 0.407 |
| Employed | -0.009 | -0.309 | 0.292 | 0.955 |
| Type of participant's family |  |  |  |  |
| (Constant) | 0.464 | 0.248 | 0.680 | 0.001 |
| Nuclear | -0.128 | -0.374 | 0.118 | 0.308 |
| Extended | 0.128 | -0.118 | 0.374 | 0.308 |
| History of SCD in participant's family |  |  |  |  |
| (Constant) | 0.377 | 0.264 | 0.489 | 0.001 |
| Yes | -0.072 | -0.354 | 0.211 | 0.619 |
| No | 0.072 | -0.211 | 0.354 | 0.619 |

### Multiple linear Regression Results on the Level of Knowledge Change toward Sickle Cell Disease among University Students

Multiple linear regressions analysis model of knowledge against predictors such as use of media televion, history of SCD in participants ‘family was performed. The results indicated knowledge change was not significant associated with other predictors (p>0.05). (Table 5)

**Table 5.** Multiple linear Regression Results on the Level of Knowledge Change on Sickle Cell Disease among University Students (n=697)

| Variable | Knowledge Change |  |  |  |
| --- | --- | --- | --- | --- |
|  | B | 95% CI |  | P-Value |
|  |  | Lower | Upper |  |
| (Constant) | 0.187 | -0.149 | 0.523 | 0.275 |
| Zones of participants |  |  |  |  |
| Coastal Zone | -0.003 | -0.323 | 0.317 | 0.985 |
| Northern Zone | 0.096 | -0.158 | 0.349 | 0.458 |
| Southern Highland Zone | 0.017 | -0.280 | 0.314 | 0.909 |
| Media uses by participants |  |  |  |  |
| Television | 0.007 | -0.274 | 0.287 | 0.964 |
| Education level of participants 'father |  |  |  |  |
| College | -0.133 | -0.479 | 0.213 | 0.450 |
| Education level of participants 'mother |  |  |  |  |
| College | 0.440 | 0.033 | 0.846 | 0.034 |
| Primary education | -0.059 | -0.297 | 0.178 | 0.623 |
| Occupational status of participants 'father |  |  |  |  |
| Self employed | -0.183 | -0.476 | 0.111 | 0.222 |
| Occupational status of participants 'mother |  |  |  |  |
| Self employed | 0.321 | 0.016 | 0.626 | 0.039 |
| Type of participant's family |  |  |  |  |
| Extended | 0.138 | -0.111 | 0.387 | 0.277 |

## 4.7 Discussion of the Findings

### Distribution of Social-Demographic characteristics of the study participants

The results of the current study on respect of the participants age, indicate that majority of them were in early young adult and unmarried. This implies, during this age majority are at the university level thus we can impart and promote them important of improving health behavior on knowledge on preventing genetic inheritance disease like Sickle Cell. This result is similar to the study conducted by Mohamed *et al*. (2015), who reported that the age of students ranged between 20 and 25 years. Moreover, the study done at Tertiary Institution in Sokoto State, Nigeria by Ango *et al*. (2018), reported the most prevalent age was 20 to 24 years.

In relation to sex, a larger proportion were males as compared to female, the proportional imitation with national statistics of high education enrollment, of which more male students enroll in high education as compared to girls (TCU, 2017). This is in agreement with the study by Ango *et al*. (2018) reported 70.3% of participants were males. Contrary to this finding, a study done at Lagos state University Olatona & Odeyemi (2012), reported female to be more than half (59.1%). Although in most low and middle economical income countries males are more dominant in high educational level as compared to female, still both have equal chance to gain knowledge towards premarital Sickle Cell genotype screening.

In respect of media for communication, mobile phones were most frequently used followed by other sources such as magazines, radio, and television. This match by the study conducted by Ugwu, (2016), who reported radio, and television were (10.3%) used by students as a source for getting information. Similar findings in a cross-sectional study at Ghana by Boadu (2018) who reported media use was (12.6%) among students. This indicates that media can be an effective way of educating the community, especially university students important of screening for SCD before marriage. Moreover, special apparatus can be designed and uploaded in mobile phones which can be reminding the phone users an important of SCD screening. This could help community as whole to be informed and take appropriate action towards prevention of Sickle Cell disease.

### Overall Level of Knowledge on sickle cell disease among university students

The results of the current study showed a significant increase in posttest mean score on the level of knowledge on Sickle Cell disease among university students. The result is related to the study at Saudi Arabia by Kotb *et al*. (2019) assessing the effect of a health education programme on the knowledge of and attitude about Sickle Cell disease among male secondary school students which reported, the mean student knowledge score was 6.04 ± 3.02 on the pretest, which improved to 10.73 ± 3.47 on the posttest, with a statistically significant difference (t = 15.2, p < 0.001). Moreover, the current results correlate to those of the study by Ango *et al*. (2018b) reported the proportion of respondents with good knowledge at pretest was 55(49.54%) which increase to 97 (87.38%) at posttest after health education (p <0.05). Although there were increases in knowledge level, as compared to previous studies the sample size was different. This implies that health education has positive effects on knowledge change regardless of the number of people.

In respect to items analysis, majority of participants at pretest, were having low knowledge on most of items of Sickle Cell disease such as; domain of SCD causes, at pretest, few of them responded correctly about, when both parents have SCD trait, the chance of each pregnant having a child with Sickle Cell disease is 25%, after heath education more than three quarter responded properly. This result is correlating to findings of the study conducted by Bagudo (2019), assessing the level of knowledge on the cause and preventive of Sickle Cell disease among Primary Education Students Nigeria, where at pretest, only (22%) responded correctly on the causes of SCD, but at posttest (88%) responded correctly.

About, screening domain, at pretest more than half of the participants were incapable of items such as SCD screening is a test done before marriage to rule out Sickle Cell disorder, and a negative Sickle Cell carrier test means Personal is very unlikely to have a disease but after intervention majority. The current finding is correlated with Kotb *et al*. (2019b) who reported at pretest the screening domain was (23.2%), after health education intervention the scores raised to (86.7%). Likewise, about management domain, in an item which required knowing if people with SCD need Medicine and people with SCD need a regular blood transfusion, participants less than half at pretest was responded correctly while at posttest more than three quarters were able to respond appropriately. This finding is similar to Bagudo (2019), who reported at pretest only (33.3%) responded correctly on the management of Sickle Cell disease while at posttest (91.3%) responded correctly on the management of SCD.

Moreover, regarding of Sickle Cell disease complications domain on the items such as; people with sickle SCD are more likely to develop pain requiring hospitalization; people with sickle SCD are more likely to develop kidney failure and Stroke, less than quarter participants were able to respond correctly, after intervention at posttest more than three quarters were able to answer properly. Nevertheless, in Tanzania, though the prevalence of Sickle Cell disease is higher, there is a low level of knowledge on SCD to the community (Ambrose *et al*., 2018). This creates a need for developing curriculum from primary education to university level which introduces topics related to premarital genotype Sickle Cell screening and counselling equally to arts and science studies for the purpose of controlling the disease in the country.

### 4.8.2 Mean score differences in pre and posttest level of knowledge on sickle cell disease among university students

The findings of this study indicated a significant difference between pre and posttest levels of knowledge towards premarital genotype sickle screening among university students. Improvement of the level of knowledge was the outcome of health education. This is in agreement with the results of the previous study carried out in Saudi Arabia which found that imparting health education could significantly raise the level of participants’ knowledge concerning Sickle Cell disease (Kotb *et al*., 2019b). Similar findings with the study by Ango *et al*. (2018) reported there were mean score difference between pretest and post-test.

Furthermore, the current results are in accordance with the study conducted among Students in Sokoto State which reported the mean scores were significantly improved regarding premarital genetic screening at pre-counseling and post counseling intervention (Abioye-Kuteyi *et al*., 2018). This also matches a study conducted in Baghdad University by Salman and Abass (2019), who reported highly significant mean differences at pre and posttest toward the impact of the programme through raising knowledge grades of studied participants. The similarities to current and previous studies could be due to the same study design, study populations, and inclusion criteria for study participants. On another hand both current and previous studies insisted significance of health education on improving the level of knowledge towards premarital genotype Sickle Cell screening, the focus is to prevent inheritance of disease traits to coming offspring.

### The predictors associated with knowledge change towards Sickle Cell disease among university students

The results of current study indicate that no any predictors were associated with knowledge change. Change in knowledge was associated with Health education provided to participants. The findings is Similar to study in Saudi Arabia assessing effect of health education programme on the knowledge about sickle Cell disease among male Secondary students by Kotb et al., (2019) who reported that health education programme significantly improved the overall level of students’ knowledge of sickle cell anemia and premarital screening. The results are congruent with study by Ango et al., (2018) which reported Health education intervention was effective in improving the knowledge of SCD and practice of genotype Voluntary screening and Counseling among the participants. Moreover, similar findings reported by Abioye-Kuteyi, Oyegbade, Bello, & Osakwe, (2018). Additionally, a study at El Minia University regarding premarital Screening and Counseling aimed to improve SCD knowledge among medical and non-medical students conducted by Abedel-Azim Mohamed, (2015) reported health education improved the knowledge of both group respectively. These findings imply that education could be effective on influence of behavior changes. Regularly, informing students on sickle cell disease and important of screening before marriage brings positive outcome to community towards the disease and its preventive measures.

## 5.1 Conclusion

The current study has tested the efficacy of premarital genotype screening and counselling on knowledge toward Sickle Cell disease among university students at baseline and re-assessed after applying education intervention to the same employed participants. The findings showed that there was a significant mean difference in knowledge between pre and posttest. Moreover, the results indicate scientific confirmation that predictors were not significantly associated with the change of knowledge, only health education on sickle cell disease associated with knowledge change among participants.

## 5.3 Recommendations

- There is a need for Ministry of Health, Community Development, Gender, Elderly and Children Tanzania to develop a policy which will motivate youth in the community to build an attitude of performing Sickle Cell screening before marriage to minimize transmitting Sickle Cell genetic disorder to their offspring. Whereby those with Sickle Cell trait (Hb AS) should be encouraged to marry with those who do not have sickle genotype (Hb AA).
- In all Universities Tanzania, Premarital genotype Sickle Cell screening and counselling education should be continuous and essential to all first-year students through highlighting to them the risk of having a generation of Sickle Cell in our Nation.

## 5.4 Suggestion for Further Research

- The present study was based to college students, it could be more preferably another study to be conducted among adolescents from primary and secondary school because they have not so far started a relationship as compared to college students, of which sometimes it makes difficult for them to break a relationship if found both of them have Sickle Cell Trait.
- The current study was uncontrolled quasi-experimental, the suggestion for the coming study could be with a control group to obtain the effectiveness of intervention between intervention and control groups.

## Data Availability

All relevant data are within the manuscript and its Supporting Information files.

## Aknowlegdment

The authors acknowledge Dr. Stephen M. Kibusi (Ph.D.), supervisor, all staff from The university of Dodoma School of Nursing and Public Health and students from all collegies of the university of Dodoma for their participations

## Author Contributions

A.G.L Developed and designed the proposal, as well as research training tools and study interventions. S.M.K. : Developed the study idea/concept, reviewed, assessed, and edited the work using re-search training resources and research tools.

## Ethical approval

Approval by the University of Dodoma (UDOM) Institutional Research Review Committee and agreement to participate: appropriate, approved by the University of Dodoma (UDOM) Research Ethics Committee (IRRC).Ethics clearance to enter other collegies: principals and deans of the respective collegies/schools approved.

## Informed consent

For their participation in the current study, all participants were requested for informed consent.

---

**Figure 1:**
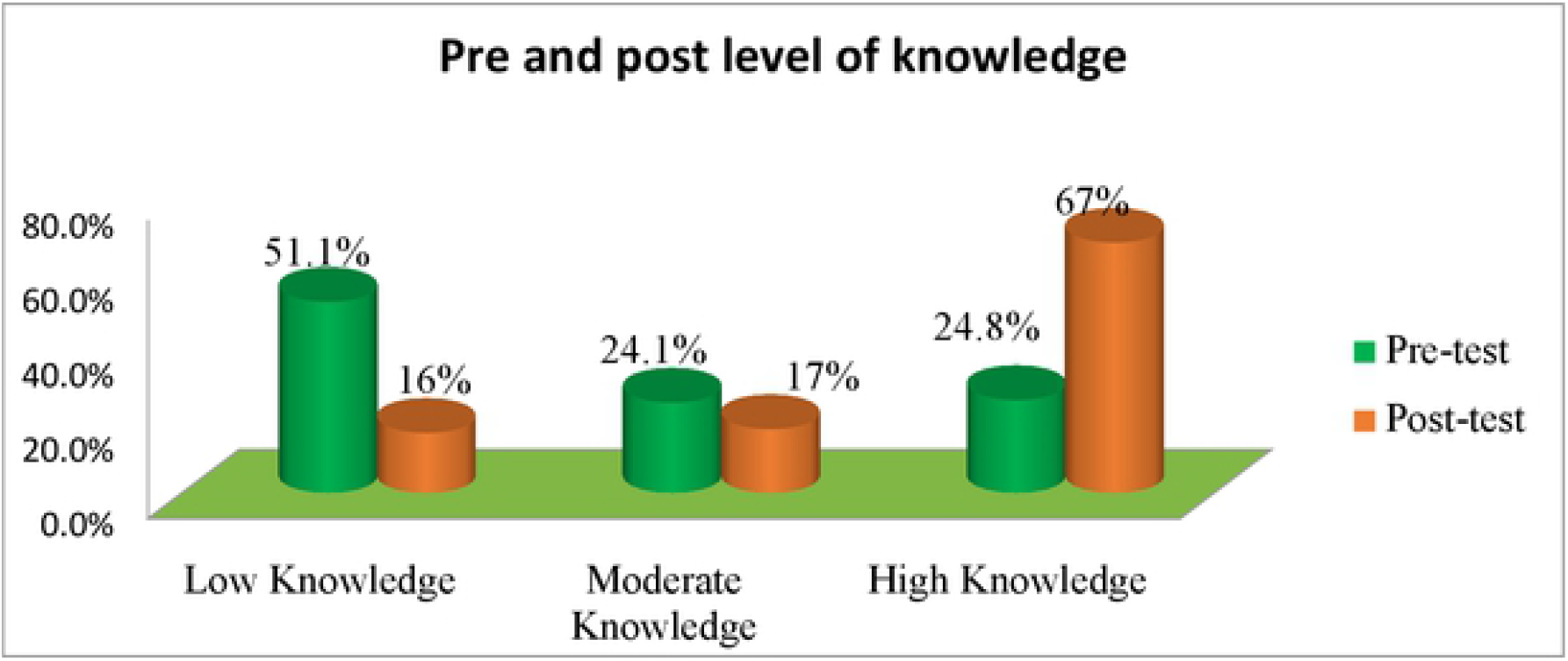
Overall pre and posttest knowledge on sickle cell disease among university students in Dodoma Region (n = 697)

## References

Abedel-Azim Mohamed, H. (2015). Improving Knowledge And Attitude Of Medical And Non-Medical Students At El Minia University Regarding Premarital Screening And Counseling. American Journal Of Nursing Science, 4(5), 270. https://Doi.Org/10.11648/J.Ajns.20150405.14

Abioye-Kuteyi, E. A., Oyegbade, O., Bello, I., & Osakwe, C. (2018). Sickle Cell Knowledge, Premarital Screening And Marital Decisions Among Local Government Workers In Ile-Ife, Nigeria. African Journal Of Primary Health Care And Family Medicine, 1(1), 53–57. https://Doi.Org/10.4102/Phcfm.V1i1.22

Aderotoye-Oni, S., Diaku-Akinwumi, I. N., Adeniran, Y., & Falase, B. (2018). Unprepared And Misinformed Parents Of Children With Sickle Cell Disease : Time To Rethink Awareness Campaigns Study Design, 10(12), 1–16. https://Doi.Org/10.7759/Cureus.3806

Al-Balushi, A. A., & Al-Hinai, B. (2018). Should Premarital Screening For Blood Disorders Be An Obligatory Measure In Oman? Sultan Qaboos University Medical Journal, 18(1), E24–E29.

Ambrose, E. E., Makani, J., Chami, N., Masoza, T., Kabyemera, R., Peck, R. N., … Smart, L. R. (2018). High Birth Prevalence Of Sickle Cell Disease In Northwestern Tanzania. Pediatric Blood And Cancer, 65(1). https://Doi.Org/10.1002/Pbc.26735

Ango, U. M., Abiola, A. O., Yakubu, A., Ibrahim, M. T. O., Awosan, K. J., & Yunusa, E. U. (2018). Effect Of Health Education Intervention On Knowledge Of Sickle Cell Disease And Practice Of Voluntary Genotype Couselling And Testing Among Students Of A Tertiary Institution In Sokoto State, Nigeria., (4), 2–7. https://Doi.Org/10.21276/Aimdr.2018.4.6.CM2

Asare, E. V, Wilson, I., Kuma, A. A. B., Dei-Adomakoh, Y., Sey, F., & Olayemi, E. (2018). Burden Of Sickle Cell Disease In Ghana : The Korle-Bu Experience. Article ID 6161270, 2018, 5.

Babalola, O. A., Chen, C. S., Brown, B. J., John, F., Falusi, A. G., & Olopade, O. I. (2019). Knowledge And Health Beliefs Assessment Of Sickle Cell Disease As A Prelude To Neonatal Screening In Ibadan, Nigeria, 3, 1–13. https://Doi.Org/10.29392/Joghr.3.E2019062

Boadu, I. (2018). Journal Of Community Medicine & Knowledge, Beliefs And Attitude Towards Sickle Cell Disease Among University Students. Journal Of Community Medicine, 8(1), 1–5. https://Doi.Org/10.4172/2161-0711.1000593

Chiao, C., Yi, C. C., & Ksobiech, K. (2012). Exploring The Relationship Between Premarital Sex And Cigarette/Alcohol Use Among College Students In Taiwan: A Cohort Study. BMC Public Health, 12(1), 1–19.

Faremi, A. F. (2018). Knowledge Of Sickle Cell Disease And Pre Marital Genotype Screening Among Students Of A Tertiary Educational Institution In South Western Nigeria Knowledge Of Sickle Cell Disease And Pre Marital Genotype Screening Among Students Of A Tertiary Educational, (October).

Faremi, A. F., & Olatubi, M. I. (2018a). Knowledge Of Sickle Cell Disease And Pre Marital Genotype Screening Among Students Of A Tertiary Educational Institution In South Western Nigeria, 11(1), 285–295.

Faremi, A. F., & Olatubi, M. I. (2018b). Knowledge Of Sickle Cell Disease And Pre Marital Genotype Screening Among Students Of A Tertiary Educational Institution In South Western Nigeria Knowledge Of Sickle Cell Disease And Pre Marital Genotype Screening Among Students Of A Tertiary Educational. International Journal Of Caring Sciences, 11(1), 285–295.

Ghimire, G. (2016). Knowledge And Attitude Regarding Sickle -Cell Disease Among Higher Secondary Students, Nepal. International Journal Of Nursing Research And Practice, 3(2), 25–30.

Hussaini, M. A., Durbunde, A. A., Jobbi, Y. D., Muhammad, I. Y., & Mansur, A. U. (2019). Assessment Of Experience, Perception And Attitude Towards Premarital Sickle Cell Disease Screening Among Students Attending Federal College Of Education, Kano, Nigeria. International Journal Of Research And Reports In Hematology, 2(February), 1–12.

Kambasu, D. M., Rujumba, J., Lekuya, H. M., Munube, D., & Mupere, E. (2019). Health-Related Quality Of Life Of Adolescents With Sickle Cell Disease In Sub-Saharan Africa : A Cross-Sectional Study, 1–9.

Kotb, M. M., Almalki, M. J., Hassan, Y., Sharif A. Al, Khan, M., & Sheikh, K. (2019). Effect Of Health Education Programme On The Knowledge Of And Attitude About Sickle Cell Anaemia Among Male Secondary School Students In The Jazan Region Of Saudi Arabia : Health Policy Implications. Biomed Research International Two, 2019, 6.

Lola, N., Kever, R. T., Uba, M., Danlami, S., Ishaku, S., Tsado, M. J., & Ibrahim, A. U. (2018). Assessment Of Knowledge Of Sickle Cell Disease And Premarital Genotyping Among Youths In Mairi Ward, Jere Local Government Of Borno State North-Eastern Nigeria. International Journal Of TROPICAL DISEASE & Health, 32(1), 1–8. https://Doi.Org/10.9734/IJTDH/2018/34442

Makani, J., Soka, D., Rwezaula, S., Krag, M., Mghamba, J., Ramaiya, K., … Disabilities, D. (2015). Health Policy For Sickle Cell Disease In Africa: Experience From Tanzania On Interventions To Reduce Under-Five Mortality. Trop Med Int Health., 20(2), 184– 187. https://Doi.Org/10.1111/Tmi.12428.Health

Makani, J., Tluway, F., Makubi, A., Soka, D., Nkya, S., Sangeda, R., … Mmbando, B. P. (2018). A Ten Year Review Of The Sickle Cell Program In Muhimbili National Hospital, Tanzania, 1–13.

Mohcdgec. (2016). Strategic And Action Plan For The Prevention And Control Of Non-Communicable Diseases In Tanzania 2020. Ministry Of Health, Community Development, Gender, Elderly And Children.

Mombo, L., Mabioko-Mbembo, G., Ontsitsagui, E., Mboui-Ondo, S., Nzé-Kamsi, L., Nkoghé, D., & Elion, J. (2017). Haemoglobin F, A2, And S Levels In Subjects With Or Without Sickle Cell Trait In South-Eastern Gabon. Hematology, 22(8), 508–513. https://Doi.Org/10.1080/10245332.2017.1292622

Omuemu, V., Obariagbon, O., & Ogboghodo, E. (2012). Awareness And Acceptability Of Premarital Screening Of Sickle Cell Disease Among Students Of The University Of Benin, Benin City. 13th World Congress On Public Health World Health Organization, (4), 1–7.

Review, A., & Bridget, O. (2015). Knowledge Attitude And Practice Towards Pre-M Aritial / Prenatal Genetic Testing Among Young People (15-45) Years Of Age In Sapele Local Government Area, Delta State. Nigeria. South American Journal Of Academic Research, 1(1), 2–38. Retrieved From https://Www.Academia.Edu/31420688

Salman, A. D., & Abass, I. M. (2019). Effectiveness Of An Instructional Program Of Premarital Screening For Hereditary Blood Diseases On Student’s Knowledge At Baghdad University. Indian Journal Of Forensic Medicine And Toxicology, 13(1), 252–258. https://Doi.Org/10.5958/0973-9130.2019.00051.3

WHO. (2017). Be Smart, Know About Sickle Cell Disease. REGIONAL OFFICE FOR AFRICA. Retrieved From https://Www.Afro.Who.Int/News/Be-Smart-Know-About-Sickle-Cell-Disease

WHO Afica, W. H. O. (2018). Be Smart, Know About Sickle Cell Disease ! Dar Es Salaam. Retrieved From https://Www.Afro.Who.Int/Health-Topics/Sickle-Cell-Disease

WHO OF, & Republic, C. (2019). Centre Of Excellence Brings Hope For Sickle Cell Patients In Congo Republic.

WORLD HEALTH ORGANISATION. (2006). Medical Genetic Services In Developing Countries The Ethical, Legal And Social Implications Of Genetic Testing And Screening. Genomics, 1–113. Retrieved From http://Www.Who.Int/Genomics/Publications/GTS-Medicalgeneticservices-Oct06.Pdf

